# Impact of Primary Graft Dysfunction on Neurodevelopmental Outcomes in Pediatric Heart Transplant Recipients

**DOI:** 10.64898/2026.03.30.26349794

**Authors:** Jorge A. Monserrate-Marrero, Mario Castro Medina, Brian Feingold, Manuel Giraldo Grueso, Kisten Rose-Felker, Richard Tang, Kei Kobayashi, Carlos Diaz-Castrillon, Katie McIntyre, Luciana Da Fonseca Da Silva, Jose P. Da Silva, Victor O Morell, Laura Seese

**Author notes:** Both authors contributed equally (Jorge A. Monserrate-Marrero, Mario Castro Medina). **Correspondence and Reprint Requests:** Laura Seese, MD, Division of Pediatric Cardiothoracic Surgery, Pittsburgh Children’s Hospital of UPMC, 4401 Penn Avenue, Pittsburgh, PA 15224.

## Abstract

**Background:** Primary graft dysfunction (PGD) remains one of the leading causes of early mortality after pediatric heart transplant (HT). While neurodevelopmental impacts of congenital heart disease (CHD) are well-characterized, the effect of PGD on long-term neurodevelopmental outcomes in pediatric HT recipients remains unknown. We sought to determine the association between PGD and neurodevelopmental outcomes in this population.

**Methods:** We performed a retrospective cohort study using the United Network for Organ Sharing (UNOS) database. All pediatric (age <18 years) isolated heart transplant recipients from 2010-2025 were included. The most recent pre- and post-transplant neurodevelopmental outcomes including cognitive delay, motor development, academic progress, and function status (stratified by age) were compared between PGD (n=434) and non-PGD groups (n=6956).

**Results:** PGD patients had significantly worse pre-transplant functional status and motor development. Post-transplant, PGD was associated with worse motor development (18.8% vs. 13.0% definite motor delay; p=0.01) and functional status in younger children (39.5% vs. 57.8% able to keep up with peers; p<0.001). Post-transplant stroke occurred 3.5 times more frequently in PGD patients (11.5% vs. 3.3%; p<0.001). Cognitive development (p=0.94) and academic progress (p=0.09) did not differ significantly. Thirty-day (7.8% vs. 1.9%) and 1-year mortality (20.3% vs. 6.4%) were significantly higher in PGD patients (both p<0.001).

**Conclusions:** This is the first study to characterize neurodevelopmental outcomes in pediatric patients undergoing HT with PGD. PGD is associated with significantly worse motor development and functional status independent of pre-transplant baseline. There is a 3.5-fold higher stroke rate providing a plausible neurological mechanism. The findings support targeted developmental surveillance recommendations and early intervention for this high-risk population.

## Introduction

Pediatric heart transplantation (HT) is the definitive treatment for end-stage heart failure in children, with one-year survival now exceeding 90% in the current era^1,2^. As outcomes have improved, clinical and research focus has increasingly shifted toward long-term morbidity, innovative strategies for organ expansion, and neurodevelopmental outcomes^1,2^. Pediatric HT recipients, particularly those with congenital heart disease (CHD), are at heightened risk for neurodevelopmental delays, with well-documented vulnerabilities in expressive language, visual-motor integration, and fine motor skills^3^. Children post-HT demonstrate IQ and visual-motor scores in the borderline-to-low average range, with CHD patients scoring significantly lower, independently associated with a more complex pre- and peri-operative transplant course^4^.

Longitudinal data confirm consistently delayed motor development in infant HT recipients, with age-dependent cognitive variability and significant decreases in mental development scores observed at 18 and 28–36 months^5^. More recent single-center data report that up to 93% of children transplanted before age 2 carry a developmental delay diagnosis, with 35% diagnosed with autism spectrum disorder, and that adverse neurodevelopmental outcomes are disproportionately common among CHD patients, with CHD independently associated with a composite outcome of death, disability, and mental delay^6^.

Primary graft dysfunction (PGD), defined by the 2014 ISHLT consensus as single or biventricular dysfunction within 24 hours of transplant in the absence of secondary causes such as rejection, pulmonary hypertension, or surgical complications, remains one of the leading causes of early mortality following pediatric HT^7^. Severe PGD occurs in approximately 4.7% of all pediatric recipients and up to 7.8% of infants, with no significant decline observed over two decades, and pre-discharge mortality reaching up to 40.6% in affected patients^8,9^. Independent risk factors include younger age, CHD, pre-transplant ECMO, and prolonged graft ischemic time. Severe PGD frequently necessitates mechanical circulatory support with veno-arterial extracorporeal membrane oxygenation (VA-ECMO), which carries its own neurodevelopmental burden^8^. A recent analysis of ECMO-bridged transplant patients demonstrated a cumulative incidence of combined motor and cognitive delay in 20% of recipients at 3 years, with postoperative stroke independently associated with delay progression and developmental delay itself predicting subsequent mortality^10^. The low cardiac output state, cerebral hypoperfusion, systemic inflammatory cascade, and prolonged critical illness inherent to PGD and its management collectively create a neurologically vulnerable window. Given that even uncomplicated pediatric HT carries substantial neurodevelopmental risk, it stands to reason that PGD with its compounding hemodynamic and inflammatory insults may meaningfully worsen long-term developmental trajectories in an already vulnerable population.

Despite these parallel bodies of evidence, no study has examined the specific impact of PGD on neurodevelopmental outcomes in pediatric HT recipients. This gap is increasingly relevant as donation after circulatory death (DCD) expands in pediatric transplantation, with emerging data suggesting higher rates of PGD in this context^11–16^. Early identification of high-risk patients and timely neurodevelopmental intervention are critical, yet without data characterizing the specific developmental burden conferred by PGD, targeted surveillance recommendations cannot be established. We hypothesized that PGD is associated with worse neurodevelopmental outcomes in pediatric HT recipients and performed the first large-scale analysis of this question using the United Network for Organ Sharing (UNOS) database.

## Methods

### Study Design and Data Source

We performed a retrospective cohort study using the UNOS database, which maintains comprehensive data on all solid organ transplants in the United States, including pre-transplant demographics, operative variables, post-operative complications, and longitudinal follow-up data including neurodevelopmental assessments. This study was exempt from institutional review board approval as it utilized de-identified registry data and focused on quality improvement.

### Study Population

We included all isolated heart transplant recipients (age <18 years) transplanted between 2010-2025. Exclusion criteria included multi-organ transplantation, combined lung transplants, and death within 24 hours of transplant. The final cohort comprised 7,390 patients.

### Definition of Primary Graft Dysfunction

PGD was defined using UNOS-coded variables, including reported PGD at 24 hours and/or 72 hours (left ventricular, right ventricular, and biventricular inclusive) or the requirement of postoperative extracorporeal membrane oxygenation (ECMO) at 24 hours and/or 72 hours, consistent with the ISHLT framework^7^. Patients meeting any of these criteria were classified in the PGD (n=434) cohort. All others comprised the non-PGD group (n=6,956).

### Neurodevelopmental Outcome Variables

UNOS collects clinician-reported neurodevelopmental assessments and listings at follow-up. Domains analyzed included cognitive development (no delay, questionable, probable, or definite delay), motor development (no delay, questionable, probable, or definite delay), academic progress (within grade level, delayed grade level, special education), and functional status using age-appropriate scales: a Karnofsky-based scale for adolescents and a Lansky-based scale for younger children.

### Statistical Analysis

Data are presented as means and standard deviations (SD) for Gaussian continuous variables, medians and interquartile ranges (IQR) for non-Gaussian continuous variables, and as frequencies for categorical variables. Student’s t-tests, Wilcoxon-Mann-Whitney tests, and χ2 or Fisher’s exact tests were utilized. Patients contributed follow-up time from HT until the first event or last date of contact with the UNOS registry. All tests were two-sided, with an alpha level of 0.05 used to determine statistical significance. The statistical analyses were performed using version 19 STATA software (StataCorp LP, College Station, TX, USA).

## Results

### Study Population and Baseline Characteristics

A total of 7,390 pediatric HT recipients were included, of whom 434 (5.9%) met criteria for PGD, with a median follow-up of 4.9 years (IQR 1.8–8.8) from transplant to the most recent neurodevelopmental assessment. Baseline demographics are shown in Table 1. The two groups were well-matched for age, gender, race, and blood type (all p>0.05). However, PGD patients had significantly worse pre-transplant clinical status: among adolescents, 20.5% were hospitalized versus 6.6% in the non-PGD group (p<0.001), and among younger children who were developmentally expected to be active, 31.6% were bedbound versus 8.4% (p<0.001) in the non-PGD group. Pre-transplant motor development was significantly worse in PGD patients (p<0.001), with only 46.4% having no delay versus 59.6%; cognitive development and academic progress did not differ significantly at baseline.

**Table 1:**
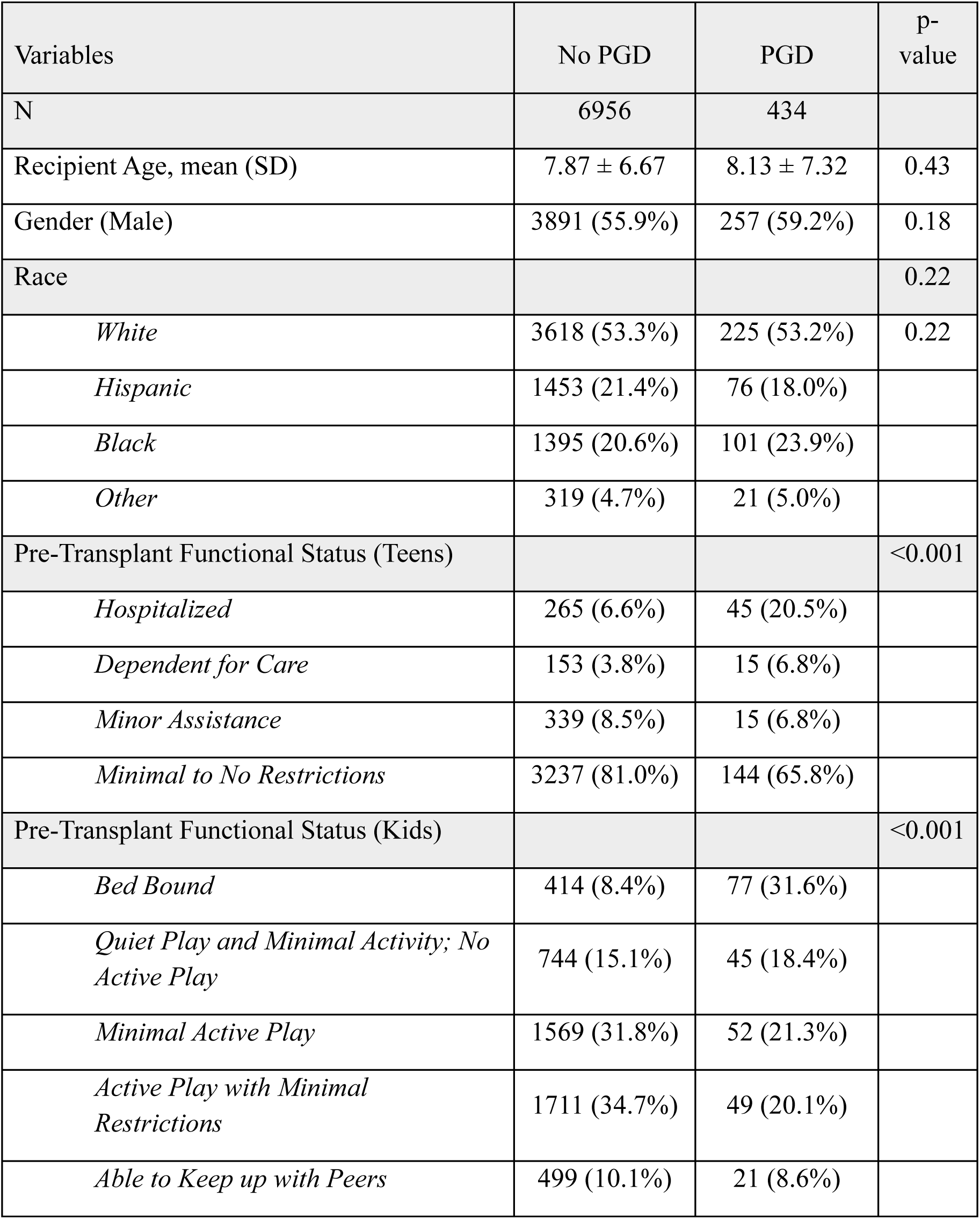
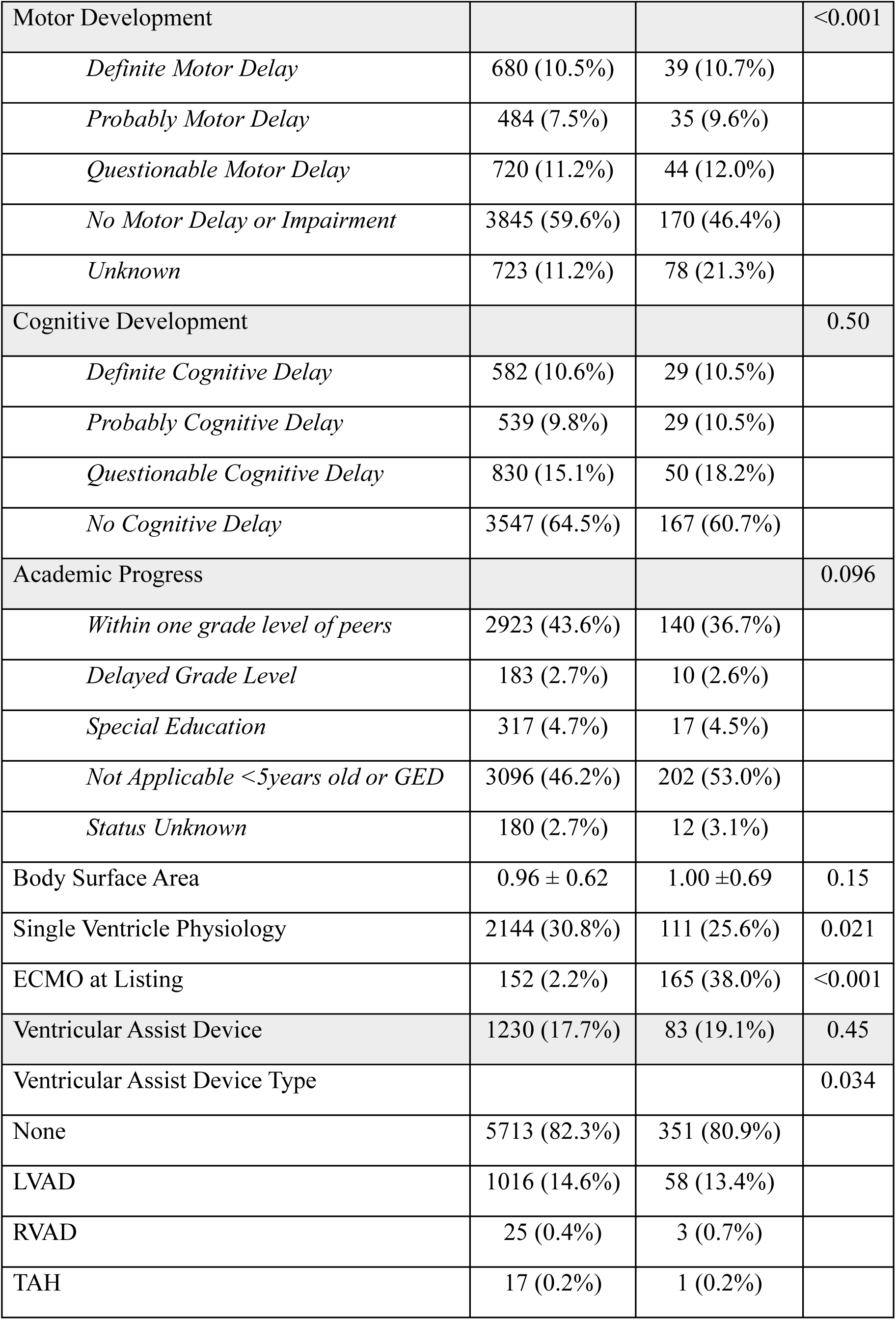
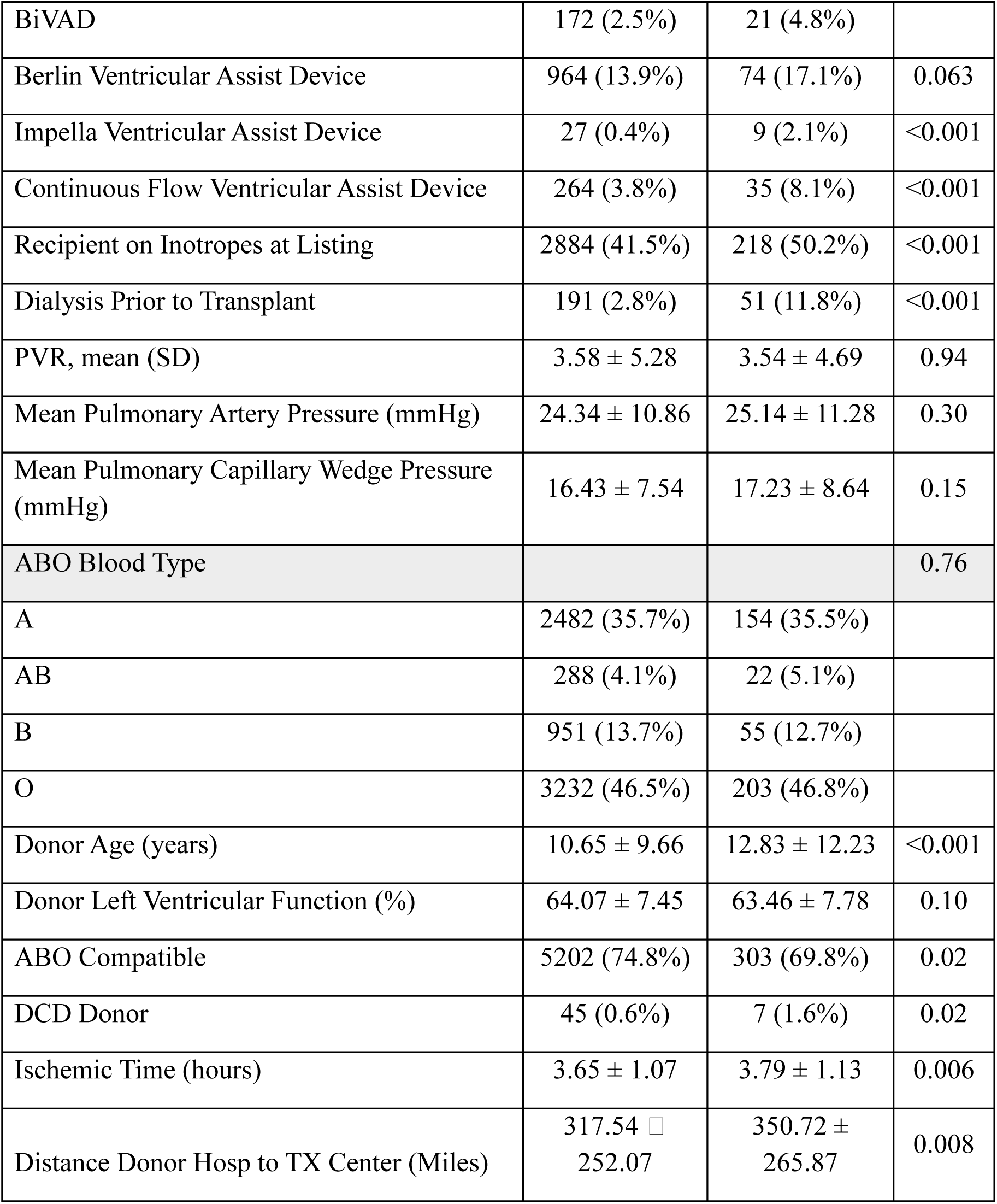
Recipient Pre-Transplant Demographics and Clinical Status.

PGD patients had substantially higher rates of pre-transplant ECMO (38% vs. 2.2%), inotrope use (50.2% vs. 41.5%), and dialysis (11.8% vs. 2.8%, all p<0.001). Single-ventricle physiology was less common in the PGD group (25.6% vs. 30.8%, p=0.021). Donor and transplant characteristics also differed, with PGD patients having older donors (12.8 vs. 10.7 years), longer ischemic times (3.8 vs. 3.7 hours), greater procurement distances (350.7 vs. 317.5 miles), and higher rates of DCD donors (1.6% vs. 0.6%, all p<0.05).

### Postoperative Outcomes

Postoperative complications are detailed in Table 2. The vast majority of PGD patients (90.3%) required post-transplant ECMO, with 51.7% on support at 24 hours and 40.3% still requiring it at 72 hours. PGD was associated with a 3.5-fold higher rate of post-transplant stroke (11.5% vs. 3.3%), more than 3-fold higher rates of renal failure requiring dialysis (21.4% vs. 6.5%), and more than twice the rate of infections treated with antibiotics (29.4% vs. 13.1%, all p<0.001). Length of stay was longer (50.1 vs. 32.2 days, p<0.001) for PGD recipients. Thirty-day mortality (7.8% vs. 1.9%) and 1-year mortality (20.3% vs. 6.4%) were both significantly higher in PGD patients (p<0.001). Notably, rejection within 1 year was less frequent in the PGD group (11.1% vs. 14.7%, p=0.036).

**Table 2:** Post-Operative Events.

| Variables | No PGD | PGD | p-value |
| --- | --- | --- | --- |
| N | 6956 | 434 |  |
| Post-Transplant ECMO at any time period | *Provided for reference. All Values 0% as per study design.* | 392 (90.3%) |  |
| Post-Transplant ECMO at 24 hours |  | 92 (51.7%) |  |
| Post-Transplant ECMO at 72 hours |  | 72 (40.4%) |  |
| Post-Transplant Complications |  |  |  |
| Stroke | 228 (3.3%) | 49 (11.5%) | <0.001 |
| Rejection Episode within 1-Year of Transplant | 1024 (14.7%) | 48 (11.1%) | 0.036 |
| Renal Failure Requiring Dialysis | 454 (6.5%) | 93 (21.4%) | <0.001 |
| Infection Requiring Antibiotic Treatment | 899 (13.1%) | 126 (29.4%) | <0.001 |
| Pacemaker Placement | 49 (0.7%) | 4 (0.9%) | 0.61 |
| Mortality at 30-Days | 134 (1.9%) | 34 (7.8%) | <0.001 |
| Mortality at 1-Year | 442 (6.4%) | 88 (20.3%) | <0.001 |
| Length of Stay Post-Transplant | 32.22 ± 39.44 | 50.10 ± 44.96 | <0.001 |

### Post-transplant Neurodevelopmental Outcomes

Post-transplant neurodevelopmental outcomes are presented in Table 3 and Figures 1–2. Across multiple developmental domains, PGD patients consistently demonstrated worse outcomes compared to non-PGD patients, with the most striking differences observed in motor development and functional status in younger children.

**Figure 1:**
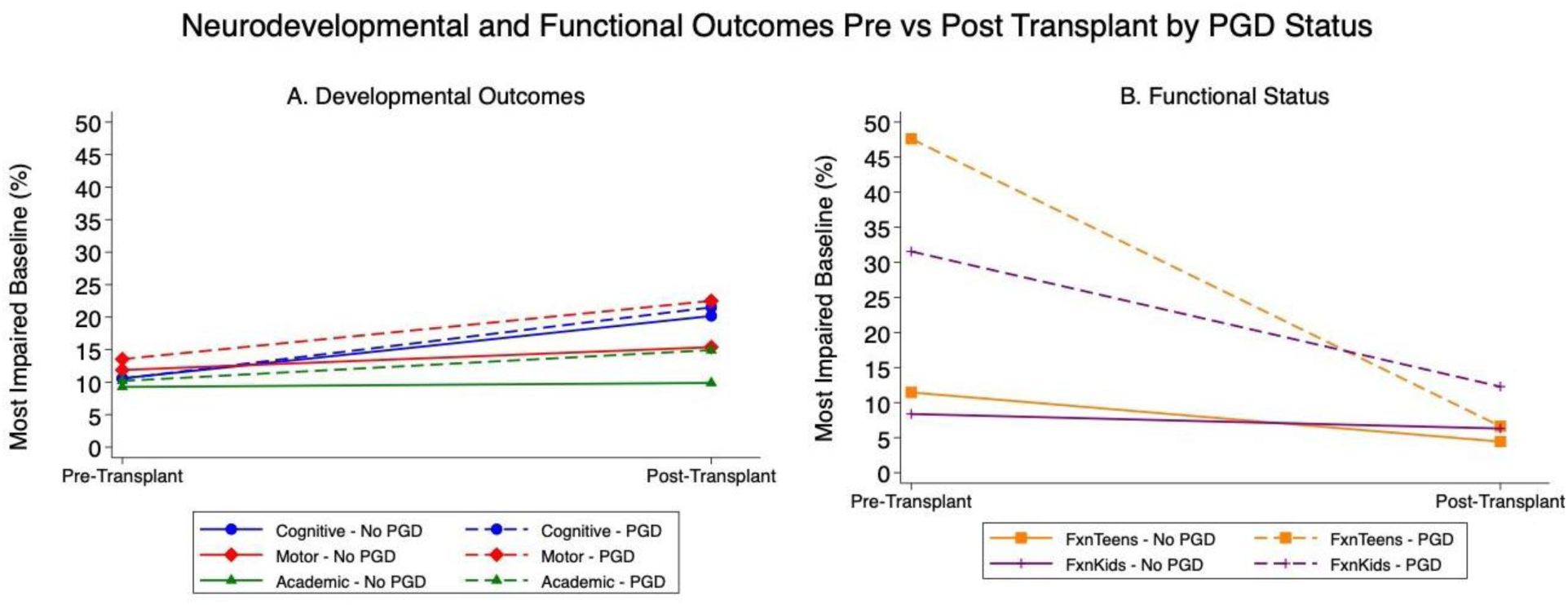
Neurodevelopmental and Functional Outcomes Pre-vs. Post-Transplant by PGD Status: Most Impaired Baseline Patients. **Panel A** shows developmental outcomes (cognitive, motor, academic); **Panel B** shows functional status by age group. Solid lines = No PGD.

**Figure 2:**
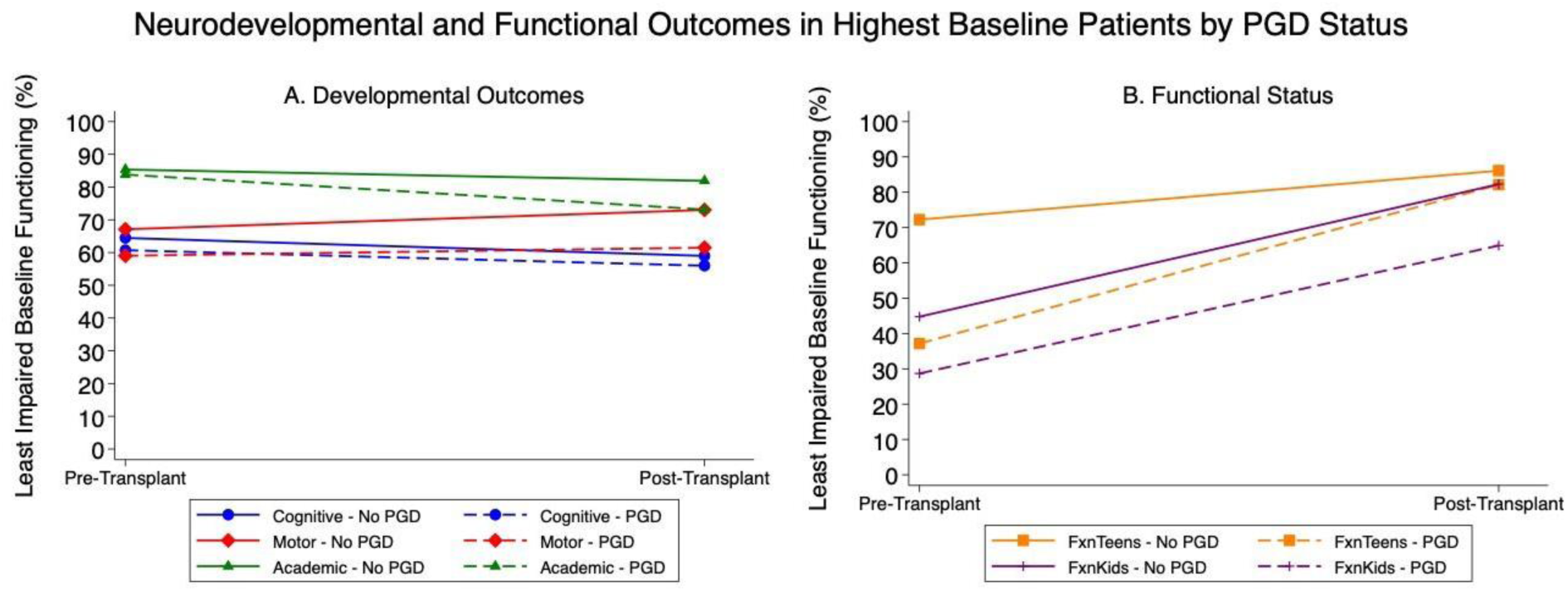
Neurodevelopmental and Functional Outcomes in Highest Baseline (Least Impaired) Patients by PGD Status. **Panel A** shows developmental outcomes; **Panel B** shows functional status. Solid lines = No PGD; dashed lines = PGD.

**Table 3:**
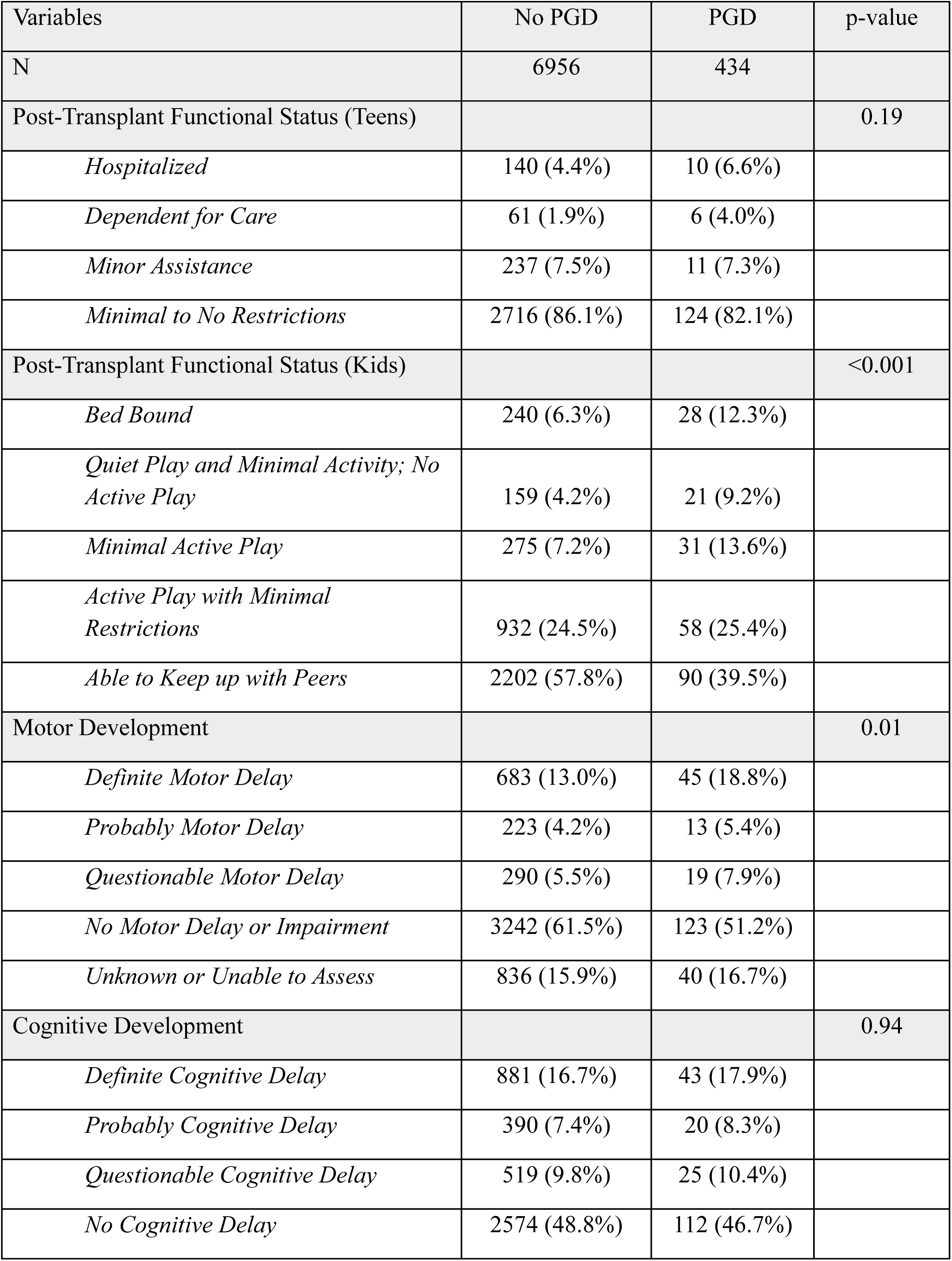
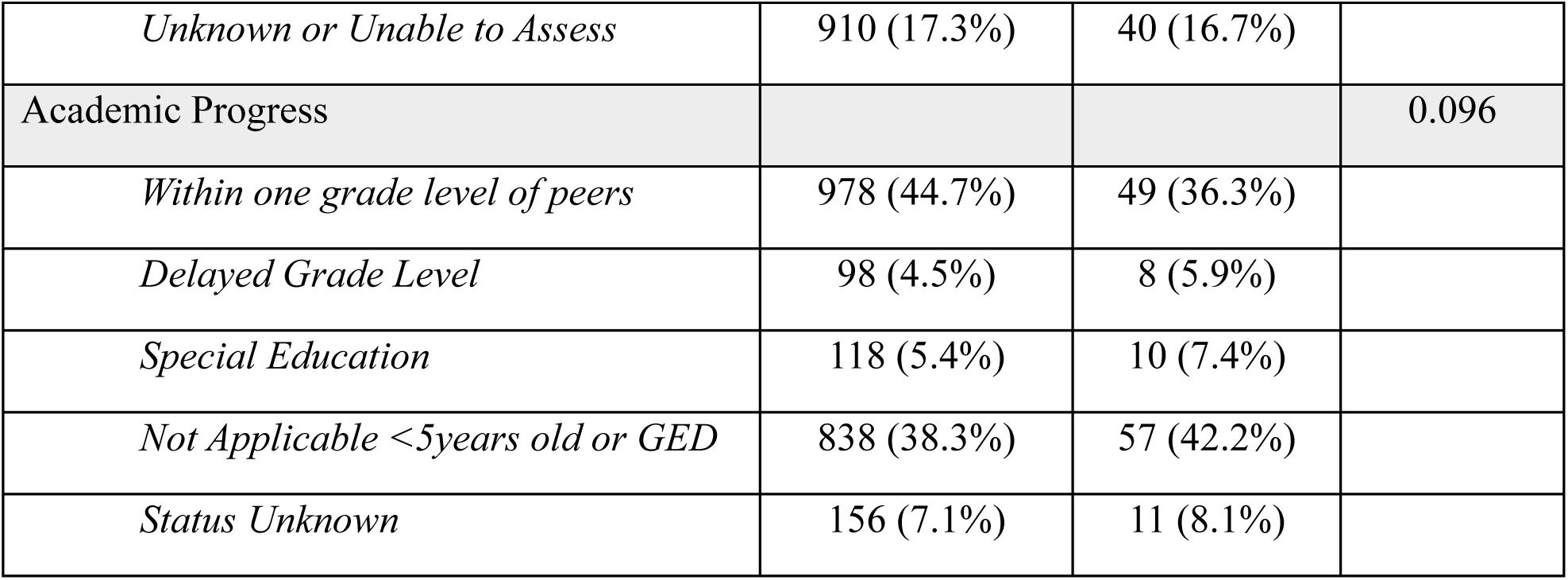
Post-Transplant Developmental Outcomes.

| Variables | No PGD | PGD | p-value |
| --- | --- | --- | --- |
| N | 6956 | 434 |  |
| Post-Transplant Functional Status (Teens) |  |  | 0.19 |
| *Hospitalized* | 140 (4.4%) | 10 (6.6%) |  |
| *Dependent for Care* | 61 (1.9%) | 6 (4.0%) |  |
| *Minor Assistance* | 237 (7.5%) | 11 (7.3%) |  |
| *Minimal to No Restrictions* | 2716 (86.1%) | 124 (82.1%) |  |
| Post-Transplant Functional Status (Kids) |  |  | <0.001 |
| *Bed Bound* | 240 (6.3%) | 28 (12.3%) |  |
| *Quiet Play and Minimal Activity; No Active Play* | 159 (4.2%) | 21 (9.2%) |  |
| *Minimal Active Play* | 275 (7.2%) | 31 (13.6%) |  |
| *Active Play with Minimal Restrictions* | 932 (24.5%) | 58 (25.4%) |  |
| *Able to Keep up with Peers* | 2202 (57.8%) | 90 (39.5%) |  |
| Motor Development |  |  | 0.01 |
| *Definite Motor Delay* | 683 (13.0%) | 45 (18.8%) |  |
| *Probably Motor Delay* | 223 (4.2%) | 13 (5.4%) |  |
| *Questionable Motor Delay* | 290 (5.5%) | 19 (7.9%) |  |
| *No Motor Delay or Impairment* | 3242 (61.5%) | 123 (51.2%) |  |
| *Unknown or Unable to Assess* | 836 (15.9%) | 40 (16.7%) |  |
| Cognitive Development |  |  | 0.94 |
| *Definite Cognitive Delay* | 881 (16.7%) | 43 (17.9%) |  |
| *Probably Cognitive Delay* | 390 (7.4%) | 20 (8.3%) |  |
| *Questionable Cognitive Delay* | 519 (9.8%) | 25 (10.4%) |  |
| *No Cognitive Delay* | 2574 (48.8%) | 112 (46.7%) |  |
| *Unknown or Unable to Assess* | 910 (17.3%) | 40 (16.7%) |  |
| Academic Progress |  |  | 0.096 |
| *Within one grade level of peers* | 978 (44.7%) | 49 (36.3%) |  |
| *Delayed Grade Level* | 98 (4.5%) | 8 (5.9%) |  |
| *Special Education* | 118 (5.4%) | 10 (7.4%) |  |
| *Not Applicable <5years old or GED* | 838 (38.3%) | 57 (42.2%) |  |
| *Status Unknown* | 156 (7.1%) | 11 (8.1%) |  |

### Motor Development

PGD patients demonstrated significantly worse post-transplant motor development (p=0.01). Only 51.2% of PGD patients had no motor delay post-transplant, compared to 61.5% in the non-PGD group. At the more severe end of the spectrum, 18.8% of PGD patients had definite motor delay versus 13.0% in the non-PGD group. These differences persisted after accounting for the significantly worse pre-transplant motor development baseline in the PGD group, suggesting that PGD confers an independent adverse effect on motor trajectories beyond what would be expected from pre-transplant disease burden alone.

### Functional Status in Younger Children

Functional status demonstrated the most clinically significant differences between groups, particularly among younger children. Post-transplant, only 39.5% of PGD patients in this age group were able to keep up with peers in typical daily activities, compared to 57.8% in the non-PGD group (p<0.001), which is a difference of nearly 20 percentage points. At the more severe end, 12.3% of PGD children were bedbound post-transplant versus 6.3% in the non-PGD group, representing a nearly two-fold difference. These findings are particularly notable given that functional status improved post-transplant in both groups overall, suggesting that while transplantation provides meaningful functional benefit, PGD substantially attenuates this recovery in young children.

### Functional Status in Adolescents

Among adolescent recipients, functional status differences between PGD and non-PGD patients did not reach statistical significance (p=0.19). However, directional trends were consistent with worse outcomes in the PGD group, including higher rates of hospitalization (6.6% vs. 4.4%) and care dependency (4.0% vs. 1.9%). The lack of statistical significance in this age group may reflect smaller sample sizes, the greater physiologic reserve of older children, or differential sensitivity of the functional status assessment tool across age groups.

### Cognitive Development

Despite substantially higher rates of ECMO exposure and post-transplant stroke in the PGD group, formal cognitive assessments did not differ significantly between groups post-transplant (p=0.94). This null finding warrants careful interpretation. It may reflect true domain-specific resilience, whereby the developing brain demonstrates relative sparing of cognitive pathways even in the context of significant hemodynamic and neurological insult. Alternatively, it may reflect the inherent limitations of the UNOS cognitive assessment tool, which relies on clinician-reported categorical data rather than standardized neuropsychological testing, potentially lacking the sensitivity to detect subtle but clinically meaningful differences. Longitudinal neuropsychological follow-up studies with validated instruments will be essential to more definitively characterize cognitive trajectories in this population.

### Academic Progress

Academic progress showed a consistent directional trend toward worse outcomes in the PGD group, with only 36.3% of PGD patients performing at grade level compared to 44.7% in the non-PGD group, though this difference did not reach statistical significance (p=0.096). The trend is nonetheless clinically meaningful. This trend, combined with the significant motor and functional status findings, supports the hypothesis that PGD has broad developmental consequences that extend into the academic domain long-term, and warrants prospective evaluation with more granular outcome measures.

### Pre-to Post-Transplant Trajectories

As illustrated in Figure 1, examining the trajectory of neurodevelopmental outcomes from pre-to post-transplant provides important context for interpreting the between-group differences observed at each timepoint. Notably, the proportion of patients in the most impaired category increased from pre-to post-transplant across developmental domains in both groups, a finding that likely reflects the cumulative neurodevelopmental burden of critical illness, prolonged hospitalization, and the physiologic stress of transplantation itself rather than a treatment-related deterioration. Across all domains, PGD patients demonstrated consistently higher rates of impairment at both timepoints, underscoring that the neurodevelopmental disadvantage associated with PGD is not simply a reflection of worse pre-transplant status but persists and in some domains widens following transplantation.

Functional status (Figure 1, Panel B) demonstrated a distinct pattern relative to other developmental domains. Unlike motor and cognitive outcomes, functional status improved post-transplant in both groups, consistent with the expected restoration of cardiac output and physical capacity following successful transplantation. However, PGD patients started at a lower functional baseline and remained lower post-transplant, with the gap between groups most pronounced among younger children. This suggests that while transplantation provides meaningful functional benefit even in the setting of PGD, the magnitude of recovery is substantially attenuated, and these children fail to achieve the same degree of functional normalization as their non-PGD counterparts.

Figure 2 further refines this picture by stratifying patients according to baseline developmental status. Among patients with the least impaired pre-transplant baseline, both PGD and non-PGD groups showed broadly similar developmental trajectories across cognitive and motor domains, suggesting that when pre-transplant reserve is preserved, the incremental impact of PGD on these specific outcomes may be less pronounced. However, even within this lower-risk subgroup, PGD patients remained consistently worse in functional status at both timepoints, reinforcing that functional impairment represents a particularly robust and persistent signal of PGD’s neurodevelopmental impact. Taken together, these trajectory analyses suggest that PGD does not simply shift the entire developmental distribution downward uniformly but rather exerts its most durable effects on motor development and functional status, which are domains that are closely tied to the physical and neurological consequences of prolonged low cardiac output and mechanical circulatory support.

## Discussion

This study is the first to characterize neurodevelopmental outcomes in pediatric heart transplant recipients with primary graft dysfunction, addressing a critical gap in the literature. Using a nationally representative cohort of 7,390 pediatric HT recipients, we demonstrated that PGD, which affects a discrete, identifiable subgroup of 434 patients (5.9%), is associated with significantly worse motor development and functional status in younger children, with a 3.5-fold higher stroke rate providing a plausible neurological mechanism. Because PGD affects a relatively small and well-defined proportion of the transplant population, it represents a high-yield target for focused neurodevelopmental surveillance and early intervention. The findings of this study make a compelling case that PGD survivors should be flagged at the time of transplant for expedited, structured developmental follow-up rather than managed as routine post-transplant patients.

### PGD and Motor/Functional Outcomes

The significant associations between PGD and worse motor development (p=0.01) and functional status in younger children (p<0.001) are the central findings of this study. Motor development was the domain most consistently affected, well-aligned with the broader ECMO and mechanical circulatory support literature in which motor domains are recognized as particularly vulnerable^17^. Given that 90.3% of PGD patients in our cohort required post-transplant ECMO, these findings are mechanistically consistent. Critically, motor development is a domain amenable to early intervention, as physical and occupational therapy are recommended for high-risk children with CHD, and emerging evidence suggests potential benefit in motor outcomes following structured early intervention programs^18^. The motor deficits identified in PGD survivors therefore represent not only a prognostic signal but a concrete, modifiable target for early rehabilitation, and the relatively small size of the PGD population makes systematic early referral both feasible and impactful.

The age-dependent pattern of functional impairment, that is significant in younger children but not adolescents, mirrors the broader literature demonstrating that the developing brain in early childhood is especially vulnerable to hemodynamic insult. James et al. demonstrated that 93% of children transplanted before age 2 years had neurodevelopmental delays^6^, Freier et al. showed consistently delayed motor development in infant HT recipients^5^, and Joffe et al. reported CHD independently associated with a composite outcome of death, disability, and mental delay in recipients under age 6 years^19^. Singh et al. further demonstrated that PGD in infants carries distinct longer-term consequences, including continued attrition of survivors well beyond the early post-transplant period^9^. Together, these data reinforce that the youngest PGD survivors face a compounding developmental burden and represent the highest-priority subgroup for early and sustained neurodevelopmental intervention.

### The Role of Stroke

The 3.5-fold increase in post-transplant stroke in PGD patients (11.5% vs. 3.3%, p<0.001) provides a compelling and obvious neurological substrate for the motor and functional differences identified in this study. In prior large-scale analyses, postoperative stroke was independently associated with neurodevelopmental delay progression and predicted subsequent mortality, supporting stroke as a key mechanistic link in the causal pathway from PGD to neurodevelopmental impairment^10^. Early identification of stroke in PGD patients and prompt referral to neurorehabilitation services may represent one of the most actionable clinical intervention points available in the acute management period.

### Cognitive Development: An Informative Null Finding

The absence of significant cognitive differences (p=0.94) should not be interpreted as true cognitive equivalence. Studies utilizing formal psychometric testing have consistently demonstrated cognitive impairment in pediatric HT recipients, with mean IQ scores in the 86–88 range, 96% of formally tested CHD recipients demonstrating delay in at least one domain, and adaptive skills falling in the low-average to below-average range^3,4,6^. The UNOS cognitive assessment tool relies on clinician-reported categorical data rather than validated neuropsychological instruments, and almost certainly lacks the sensitivity to detect subtle but clinically meaningful differences. The trend toward worse academic outcomes in the PGD group (36.3% vs. 44.7% at grade level, p=0.096) further supports the likelihood that a true educational impact exists that this registry was underpowered to detect.

### Clinical Implications

PGD affects fewer than 6% of pediatric HT recipients, making targeted surveillance not only justified but feasible. A history of PGD should be formally incorporated into neurodevelopmental risk stratification frameworks for pediatric HT. PGD survivors should be enrolled in structured developmental surveillance programs beginning in the early post-transplant period, with early referral for physical therapy, occupational therapy, and developmental pediatrics ideally initiated prior to hospital discharge. With fewer than 450 patients affected annually in the UNOS registry, systematic early referral represents a tractable, high-yield intervention target that is entirely within reach of transplant programs committed to optimizing long-term outcomes.

As DCD donor utilization expands, the neurodevelopmental consequences of PGD documented here must be actively monitored in this evolving population. Simultaneously, PGD prevention through advances in donor organ preservation, including controlled hypothermic perfusion, normothermic ex-vivo perfusion platforms, and pre-implant risk stratification tools, remains the most effective long-term strategy^11,15,20,21^. Until prevention strategies mature, the burden falls on the clinical team to recognize PGD survivors as a distinct high-risk neurodevelopmental population and respond early, systematically, and with the same urgency applied to their cardiac recovery.

### Limitations

Several limitations should be acknowledged. Survivorship bias is inherent: PGD patients with the worst neurological injury may not have reached stability to participate in neurodevelopmental assessments, and our results may underestimate the true neurodevelopmental burden. UNOS neurodevelopmental assessments are clinical-reported categorical variables, not validated psychometric instruments, limiting sensitivity to subtle differences. Assessments were also not performed at standardized time points (median 4.9 years, IQR 1.8-8.8). The registry also does not capture ECMO duration, neuroimaging findings, genetic diagnosis, socioeconomic status, or access to early intervention services, all of which are important potential confounders. While nationally representative to US centers, findings may not generalize to other healthcare systems.

### Conclusions

In the first large-scale analysis of neurodevelopmental outcomes in pediatric HT recipients with PGD, we demonstrated that PGD is associated with significantly worse motor development and functional status, particularly in younger children. A 3.5-fold higher post-transplant stroke rate provides a plausible neurological mechanism underlying these differences. Critically, PGD affects a small and identifiable subgroup, fewer than 6% of recipients, making this a tractable, high-yield population for targeted intervention. These findings support the formal incorporation of PGD history into neurodevelopmental risk stratification frameworks and the initiation of structured developmental surveillance and early rehabilitation beginning at the time of transplant admission. Prospective studies using validated neuropsychological instruments are needed to fully characterize the cognitive and academic burden of PGD and evaluate the impact of targeted early intervention strategies on long-term developmental trajectories.

## Data Availability

All data published in the article can be shared upon request.

## Acknowledgements

None

